# EFFECTS OF HIV INFECTION AND FORMER COCAINE DEPENDENCE ON NEUROANATOMICAL MEASURES AND NEUROCOGNITIVE PERFORMANCE

**DOI:** 10.1101/2022.03.05.22271945

**Authors:** Kathryn-Mary Wakim, Edward G. Freedman, Madalina E. Tivarus, Zachary Christensen, Sophie Molholm, John J. Foxe

## Abstract

Evidence from animal research, postmortem analyses, and MRI investigations indicate substantial morphological alteration in brain structure as a function of HIV or cocaine dependence (CD). Although previous research on HIV+ active cocaine users suggests the presence of deleterious morphological effects in excess of either condition alone, a yet unexplored question is whether there is a similar deleterious interaction in HIV+ individuals with CD who are currently abstinent. To this end, the combinatorial effects of HIV and CD history on regional brain volume, cortical thickness, and neurocognitive performance was examined across four groups of participants: healthy controls, HIV-negative individuals with a history of CD, HIV+ individuals with no history of CD, HIV+ individuals with a history of CD. Our analyses revealed no statistical evidence of an interaction between both conditions on brain morphometry and neurocognitive performance. While descriptively, individuals with comorbid HIV and a history of CD exhibited the lowest neurocognitive performance scores, using Principle Component Analysis of neurocognitive testing data, HIV was identified as a primary driver of neurocognitive impairment. Higher caudate volume was evident in CD+ participants relative to CD-participants. Taken together, these data provide evidence of independent effects of HIV and CD history on brain morphometry and neurocognitive performance in cocaine-abstinent individuals.

## 1. Introduction

Even in the post-antiretroviral therapy era, only half of HIV-1 patients in the United States have achieved an undetectable viral load (HIV.gov, 2017). At every stage in the HIV epidemic, substance use remains a powerful co-factor: potentiating transmission (Timpson et al., 2003), contributing to delayed treatment onset (Girardi et al., 2004), and leading to reduced antiretroviral treatment adherence in HIV-infected patients (HIV+) (Meade et al., 2011). HIV-1 proteins and psychostimulant drugs have been shown to interact using both additive and synergistic mechanisms to promote poor neurocognitive health outcomes that complicate addiction recovery. The aim of this study is to elucidate the effect of comorbid HIV infection and CD on regional brain volume, cortical thickness, and neurocognitive function in cocaine-abstinent individuals in recovery.

Early autopsy studies of HIV+ patients have consistently demonstrated HIV-related morphological alterations in subcortical regions (Budka et al., 1987; De la Monte et al., 1987; Wiley et al., 1986), with dopamine (DA)-rich structures appearing particularly vulnerable to the effect of HIV (Aylward et al., 1993). Before antiretroviral therapy, 15-30% of patients diagnosed with acquired immunodeficiency syndrome (AIDS) developed HIV-associated dementia (Lawrence and Major, 2002; McArthur et al., 1993) characterized by extensive neurocognitive impairment (NCI) and motor slowing (Clifford and Ances, 2013). Although the advent of highly active antiretroviral therapy (HAART) has substantially reduced the frequency of HIV-associated dementia, subcortical alterations remain detectable by MRI in HIV+ patients both with and without NCI (Ances et al., 2012b; Becker et al., 2011; Correa et al., 2016). In addition, increased life expectancy of HIV patients in the HAART era is accompanied by higher rates of age-related comorbid disorders (Rosenthal and Tyor, 2019). One recent neuroimaging study showed significant gray matter atrophy in the striatum of HAART-treated, virologically controlled patients (Becker et al., 2011). This atrophy was shown to correlate with duration of infection, suggesting that progressive neurodegeneration occurs despite therapy.

Neuroimaging evidence from the post-HAART era suggests that HIV-related differences in brain macrostructural measures (e.g. subcortical volume) and microstructural measures (e.g. white matter integrity) extend into regions of cortex (Pfefferbaum et al., 2009; Thompson et al., 2005). HIV-related changes in MRI-based measures of cortical integrity have been shown in the superior (du Plessis et al., 2016; Towgood et al., 2012) and medial frontal gyrus (MacDuffie et al., 2018), corpus callosum (Ances et al., 2012b; Thompson et al., 2006), anterior cingulate cortex (ACC) (Kuper et al., 2011), orbitofrontal cortex (OFC) (Kallianpur et al., 2012; Williams et al., 2020), and insula (Kallianpur et al., 2012). Autopsies of HAART-treated HIV+ patients with HIV-associated neurocognitive disorder (HAND) show evidence of nuclear and mitochondrial DNA damage in the frontal cortex driven by oxidative stress-induced neuroinflammation (Zhang et al., 2012).

Neuroinflammation is also a major driver of neural disruption in CD. Chronic cocaine usage has been shown to rupture neurovascular capillaries in rats (Barroso-Moguel et al., 1997), leading to reduced blood brain barrier (BBB) integrity and greater permeability to pathogens, such as HIV-infected monocytes (Yao et al., 2011). Like the HIV Tat protein, cocaine has been shown to inhibit synaptic DA reuptake via direct interaction with the DA transporter protein (Midde et al., 2013). Lower estimates of cortical volume have been consistently shown in cross-sectional MRI studies comparing individuals with CD to those without CD. These regions that overlap with those showing HIV-related morphological alterations, include the orbitofrontal cortex (OFC) (Ersche et al., 2011), ACC (Alia-Klein et al., 2011; Franklin et al., 2002) (Matochik et al., 2003; Wheeler et al., 2013), and insula (Connolly et al., 2013).

Similar neurocognitive differences have been observed between chronic cocaine users and HAART-treated HIV+ patients in neurocognitive domains mediated by fronto-striatal pathways. Both groups show alterations in executive function, verbal memory, and working memory (Madoz-Gúrpide et al., 2011; Walker and Brown, 2018), although data suggest that cocaine-related NCI is attenuated following abstinence (Vonmoos et al., 2014). However, preliminary evidence indicates persistent, compounded NCI in HIV+ individuals with a history of substance dependence despite current abstinence (Martin et al., 2018), suggesting that the substance dependence in HIV+ patients can cause long-lasting neurocognitive changes which do not ameliorate along the typical abstinence-related recovery trajectory.

Previous work indicates that the combination of HIV and CD is associated with functional alteration in brain circuitry mediating high-level cognitive functions, including cognitive control (Wakim et al., 2021; Wakim et al., 2022) and decision making (Bell et al., 2020; Meade et al., 2017). The goal of this study was to examine the effects of HIV on macrostructure and neurocognitive function in cocaine-abstinent individuals with a history of CD. A 4-stage approach was employed. First, we report on results from neurocognitive testing batteries. Second, we examine macrostructural brain measures in regions that have consistently shown morphological differences as a function of HIV, CD history, and their combination--the OFC, ACC, insula, caudate, and putamen—using a region-of-interest (ROI) analysis approach. Third, an exploratory whole-brain analysis on cortical thickness and subcortical brain volume was undertaken to identify additional regions that might be implicated. Forth, we explored the relationship between cortical thickness, clinical variables (duration of cocaine abstinence, duration of HIV infection, viral load), and neurocognitive function using a whole-brain vertex-wise analysis approach.

## 2. Methods

### 2.1 Participants

MRI data from 93 participants was obtained, with data from 86 participants comprising the final MRI dataset used for analyses following quality control procedures and subject exclusions (described below). Four groups of participants were recruited: healthy controls (CD-/HIV-), HIV-individuals with a history of CD (CD+/HIV-), HIV+ individuals with no history of CD (HIV+/CD-), and HIV+ individuals with a history of cocaine dependence (CD+/HIV+). Neurocognitive testing data from all 93 participants was also obtained, with data from 90 participants comprising the dataset used for final analysis after all quality control and subject exclusions. Demographic characteristics of all participants included in analyses involving neurocognitive testing data, MRI data, or both neurocognitive and MRI data are shown in **Table 1. Table S1** shows the demographic characteristics of the subset of 86 participants in included in analyses involving MRI data following all subject exclusions. Exclusion criteria were as follows: (1) A diagnosis of current Major Depressive Disorder (MDD), Bipolar I, or Schizophrenia as assessed via the Structured Clinical Interview for the DSM-V Clinical Version (SCID-V-CV) (First, 2014), (2) A history of head injury or loss of consciousness for > 30 minutes, (3) Any self-reported history of neurological disorders or brain pathology, (4) Urine sample positive for any drug of abuse (e.g. cocaine, amphetamines, opiates) excluding marijuana/THC, (5) For CD+ participants, a minimum of two weeks of cocaine abstinence, maximum of 5 years, (6) For CD-participants, any history of drug or alcohol dependence. In the current sample, 3 CD+/HIV+ and 1 CD+/HIV-participants were excluded from analyses of MRI data due to cocaine-positive urine samples on the day of MRI scanning, and 3 participants were excluded from analyses involving neurocognitive testing measures (2 CD+/HIV+, 1 CD+/HIV-) due to cocaine-positive urine samples (n = 2) or opiate-positive urine samples (n = 1) on the day of neurocognitive testing. Because of the high prevalence of marijuana usage in the HIV+ population (Shiau et al., 2017) and the high prevalence of cigarette smoking in CD+ participants (Guydish et al., 2011), we did not exclude participants for THC+ urine samples from recent marijuana usage or for daily smoking. These measures were instead collected from all participants and compared as a function of participant group (**Table 1**). However, participants in CD-study groups (CD-/HIV+ and CD-/HIV-) were excluded for any lifetime history of any substance use disorder. For all HIV+ participants, clinical variables (CD4-count, medication regime, viral load) were gathered through medical record evaluation. This study was approved by the Research Subjects Review Board at the University of Rochester (STUDY1081). Written, informed consent was obtained from all participants in accordance with the tenets of the Declaration of Helsinki.

**Table 1:** Demographics. Sample characteristics by study group for neurocognitive assessment data analysis. N = 86 participants in this sample were included in the MRI analysis.

| n = 90 Participants | HIV- |  | HIV+ |  | Statistic | Effect Size | *p* value |
| --- | --- | --- | --- | --- | --- | --- | --- |
|  | CD- (n = 34) | CD+ (n = 21) | CD- (n = 20) | CD+ (n = 15) |  |  |  |
| Demographic Characteristics |  |  |  |  |  |  |  |
| Sex Assigned at Birth (N Male : N Female) | 13 : 21 | 13 : 9 | 13<sup>#</sup> : 7 | 8 : 7 | $\chi^2(3) = 4.382$ | $\Phi = 0.219$ | 0.223 |
| Age in years, M (SD) [range] | 42.7 (12.2) [19, 65] | 42.7 (10.0) [26, 61] | 44.7 (13.1) [23, 65] | 49.1 (7.4) [32, 58] | F(3,86) = 1.284 | $\eta^2 = 0.042$ | 0.285 |
| Race, N | | | | | $\chi^2(6) = 17.733$ | $\Phi = 0.471$ | 0.003* |
| African American | 5 | 6 | 9 | 10 |  |  |  |
| Caucasian | 23 | 16 | 10 | 5 |  |  |  |
| Asian | 6 | 0 | 1 | 0 |  |  |  |
| Education in years, M (SD) [range] | 16.5 (2.87) [12, 24] | 13.4 (2.61) [11, 23] | 13.4 (2.11) [9, 18] | 12.6 (2.17) [10, 16] | F(3,86) = 12.305 | $\eta^2 = 0.298$ | < 0.001* |
| THC positive urine, N |  |  |  |  |  |  |  |
| Date of Scan | 3 | 3 | 6 | 2 | $\chi^2(3) = 4.096$ | $\Phi = 0.223$ | 0.249 |
| Date of Cognitive Testing | 2 | 1 | 6 | 2 | $\chi^2(3) = 7.535$ | $\Phi = 0.305$ | 0.033* |
| Drug use characteristics |  |  |  |  |  |  |  |
| Current Smoker, N | 2 | 16 | 5 | 11 | $\chi^2(3) = 37.572$ | $\Phi = 0.627$ | < 0.001* |
| Lifetime cocaine usage in years, M (SD), [range] | | 15.0 (10.9) [1, 35] | | 18.5 (9.54) [2, 33] | t(34) = -1.051 | D<sup>\$</sup> = -0.355 | 0.315 |
| Duration of abstinence in years, M (SD) [range] |  | 0.974 (1.02) |  | 1.03 (1.36) | t(35) = 0.127 | D = -0.042 | 0.900 |
| KMSK, M (SD) [range] |  |  |  |  |  |  |  |
| Cocaine | 0.55 (2.02) [0, 10<sup>%</sup>] | 14.38 (1.98) [10, 16] | 0.50 (1.10) [0, 4] | 13.20 (3.26) [7, 16] | t(21.38) = 1.249<sup>&^</sup> | D = 0.457 | 0.225 |
| Opiates | 0.09 (0.514) [0, 3] | 7.68 (5.47) [0, 13] | 0 (0) [0, 0] | 1.20 (2.40) [0, 7] | t(30.819) = 4.908<sup>&^</sup> | D = 1.440 | < 0.001* |
| Tobacco | 3.16 (3.96) [0, 11] | 9.82 (3.16) [0, 13] | 5.55 (5.35) [0, 13] | 8.36 (3.15) [0, 12] | t(34) = 1.274<sup>&</sup> | D = 0.463 | 0.186 |
| Alcohol | 6.97 (3.12) [0, 12] | 11.0 (2.53) [4, 13] | 7.45 (3.42) [0, 13] | 9.60 (3.54) [0, 13] | t(35) = 1.371<sup>&</sup> | D = 0.459 | 0.179 |

| HIV Characteristics |  |  |  |  |  |  |  |
| --- | --- | --- | --- | --- | --- | --- | --- |
| HIV viral load <20 copies, N (%) | | | 20 (100%) | 13 (86.6%) | $\chi^2(1) = 2.828$ | $\Phi = -0.284$ | 0.176 |
| Current CD4 cell count, M (SD) [range] | | | 760 (589) [301, 3008] | 719 (232) [190, 1004] | $t(33) = 0.250$ | $D = 0.085$ | 0.804 |
| Years since diagnosis, M (SD) [range] | | | 16.9 (11.5) [1, 35] | 16.0 (7.61) [2, 33] | $t(32.569) = 0.216^{\wedge}$ | $D = 0.089$ | 0.784 |

<sup>#</sup> One participant listed as male has transitioned from male to female \*Statistically significant result at alpha = 0.05 &T-test between CD+/HIV+ and CD-/HIV+ ^ Estimated degrees of freedom \$D refers to Cohen's D % 1 CD-/HIV- participant who did not meet DSM-5 criteria for lifetime history cocaine dependence endorsed a one-weekend cocaine binge during late teens

### 2.2 Neurocognitive Testing

A 45-minute battery of neuropsychological tests was administered. The battery included the following measures: Wechsler Adult Test of Intelligence (WAIS) Working Memory Subsection (Arithmetic, Letter-Number Sequencing, Digit Span), Trail Making Test (Trails) subsection A and B (Reitan and Wolfson, 1995), the Hopkins Verbal Learning Test - Revised (HVLT-R) (Brandt, 1991), and the International HIV Dementia Scale (IHDS) (Sacktor et al., 2005). These assessments were chosen to assess the cognitive domains of working memory (WAIS Working Memory Subsection), processing speed (Trails A), executive function, (Trails B), learning (HVLT-R – Immediate Recall), memory (HVLT-R– Delayed Recall, HVLT-R – Recognition Discrimination, HVLT - Retention), and motor skills (IHDS).

### 2.3 Questionnaires

Participants were administered the Kreek-McHugh-Schluger-Kellogg Scale (KMSK) (Kellogg et al., 2003) and the Barratt Impulsivity assessment (BIS) (Patton et al., 1995) to assess severity of past drug usage and trait impulsivity. The KMSK is a rapid screening instrument used to quantify self-exposure to opiates, cocaine, alcohol and/or tobacco during an individual’s period of greatest consumption. The BIS is a 30-item questionnaire to assess personality traits associated with risk taking and impulsivity. BIS subsections include questions pertaining to attention impulsiveness, motor impulsiveness, and non-planning impulsiveness.

### 2.4 Magnetic Resonance Imaging

MRI was performed using a 3T whole-body Siemens Prisma scanner (Erlangen, Germany) equipped with a 64-channel head coil. High-resolution structural T1 contrast images were acquired using a magnetization prepared rapid gradient echo pulse (MPRAGE) sequence [repetition time (TR) = 2530 ms, echo time (TE) = 2.34 ms, flip angle = 7 degrees, field of view (FOV) = 256 mm, voxel size1 × 1 × 1 mm]. Data were collected as part of a larger study to assess the combined effect of HIV and CD on structural and functional brain measures (Wakim et al., 2021; Wakim et al., 2022).

### 2.5 Processing of Structural Scans

DICOM images were converted to NIFTI format using dcm2niix (Li et al., 2016). Cortical reconstruction and volumetric segmentation were performed using the Freesurfer image analysis suite v6.0 (http://surfer.nmr.mgh.harvard.edu/) (Fischl, 2012). High-resolution T1-weighted images were processed using the automated “recon-all” Freesurfer pipeline. Processing was parallelized using the GNL Parallel function in the Homebrew toolbox. Details of the recon-all Freesurfer procedure are reported in previous publications (Dale et al., 1999). Briefly, motion correction, intensity normalization, and affine Talairach transformation were performed. Following removal of extracranial signal, subcortical, gray and white matter were segmented, and pial surface boundaries were estimated. The automated cortical parcellation and cortical boundary detection were performed for each individual in standard space using Freesurfer algorithms. Full details of Freesurfer’s cortical parcellation algorithm are given in prior publications (Fischl et al., 2004).

### 2.6 Region of Interest Analysis

Ninety-four brain regions were parcellated based on the standard Desikan-Killiany atlas (Desikan et al., 2006) and extracted for each subject using the “mri_label2label” function in Freesurfer. Four regions of interest (ROIs), bilateral caudate, putamen, insula, rostral ACC, and medial OFC (**Figure 1)**, were declared *a priori* based on previous literature which indicated an effect of CD, HIV, or their combination on brain volumetrics.

**Figure 1:**
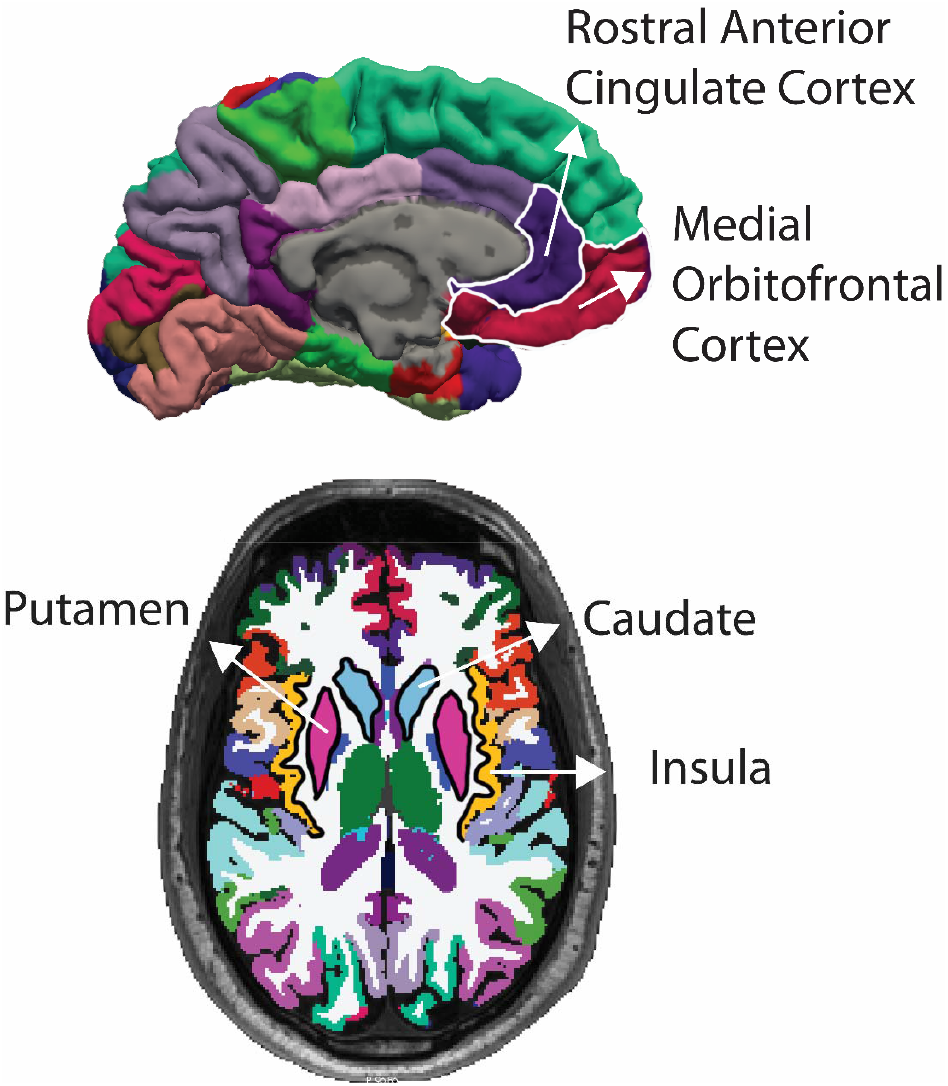
Regions of Interest: Four regions of interest encompassing the rostral anterior cingulate cortex (purple), medial orbitofrontal cortex (red), insula (gold), caudate (light blue), and putamen (pink).

### 2.7 Exploratory Vertex-Wise Analysis

To identify additional areas showing differences in cortical thickness as a function of clinical diagnosis and neurocognitive variables, a vertex-wise analysis was conducted in Freesurfer. Briefly, models of gray and white matter boundaries were constructed based on pial surface estimate computed previously during the automated “recon-all” pipeline (see section 2.5). The estimated cortical thickness measure at each point (vertex) is represented by perpendicular distance between the pial surface and the white matter boundary. Each vertex was separated was separated from neighboring vertices by approximately 1 mm, leading to total of approximately 140,000 vertices across the entire cortical surface. For each subject, cortical thickness measures were resampled to an average template in MNI space using the “mris_preproc” function (Fischl et al., 1999), allowing for vertex-wise comparisons across participants. A general linear model (GLM) was then fit to each vertex using the “mri_glmfit” function in Freesurfer (https://surfer.nmr.mgh.harvard.edu/fswiki/mri_glmfit). Group were corrected from multiple comparisons using “mri_glmfit-sim” in Freesurfer (https://surfer.nmr.mgh.harvard.edu/fswiki/mri_glmfit). A cluster-forming threshold of 2.3 (p < 0.005) was employed, and clusterwise probability was set to p < 0.05. Following correction for multiple comparisons, surviving clusters were rendered on the Freesurfer average brain (fsaverage) for visualization purposes.

### 2.8 Exploratory Analysis of Subcortical Volume and Additional Structural Measures

To examine effects of HIV, a history of CD, and the combination of these factors on subcortical structures and macrostructural measures (e.g. total brain volume), the following segmentations were extracted: nucleus accumbens, amygdala, brainstem, hippocampus, pallidum, corpus callosum, thalamus, total brain volume, total gray matter volume (including cerebellar gray matter), total cerebral white matter volume, total subcortical gray matter volume, and total ventricular volume. “Total subcortical volume” encompassed estimated volume from the bilateral caudate, putamen, thalamus, ventral diencephalon, nucleus accumbens, amygdala, hippocampus, pallidum, and substantia nigra combined.

### 2.9 Quality Control Procedures

Of 93 total scans, 4 scans (1 CD+/HIV-, 3 CD+/HIV+) were excluded from analysis due to cocaine-positive urine samples. All remaining scans were subjected to the standard Freesurfer processing pipeline. To ensure consistent quality, Freesurfer segmentations were visualized using qatools-py (https://github.com/Deep-MI/qatools-python) and manually inspected for gross segmentation errors. Qoala-T toolbox (https://github.com/Qoala-T/QC), was used to identify scans requiring further inspection and possible intervention (Klapwijk et al., 2019). Scans earning quality ratings of < 50 (n = 16 of 89 scans) using this data-driven, user-invariant approach were subjected to detailed manual inspection following the Qoala-T pipeline guidelines (https://github.com/Qoala-T/QC/blob/master/Qoala-T_ManualQC.pdf). This visual inspection procedure included rater assessments of the severity of motion artifacts, manual identification of regions containing pial surface over or underestimations, and manual verification that reconstructions did not contain non-brain tissue (e.g. skull). All scans earning a rating of “fail” by automated inspection (Qoala-T) and manual inspection (KW, ZC) were excluded (n = 3). Scans rated “poor” based on both manual and automated inspection were subjected to manual adjustment (n = 3). A total of 86 scans were included in the final analysis.

All 93 participants underwent neurocognitive testing. Of these participants, 2 CD+/HIV+ patients were excluded from analyses involving neurocognitive testing data due to cocaine-positive urine samples and 1 was excluded due to an opiate-positive urine sample. Neurocognitive data from 90 participants were included in the final analysis.

### 2.10 Statistical Analyses

Statistical analyses were conducted using IBM SPSS Statistics 27.0, 2020 and Freesurfer v.6.0.

#### 2.10.1 Demographic Data

For demographic data, 1-way ANOVAs were used to test for between-group differences among continuous variables (e.g. age, years of education). Chi-Square tests were used to identify between-group differences among categorical variables (e.g. race, sex). For analyses including categorical variables containing fewer than 5 observations per cell, Fishers Exact Test was used. Independent-sample T-tests were used when data for only two groups was present (e.g. duration of cocaine usage, duration of cocaine abstinence, viral load, CD4-count).

#### 2.10.2 Region of Interest Analysis

To examine between-group differences in macrostructural brain measures in *a priori*-defined regions of interest while controlling for the effect of demographic variables (age, sex, years of education), ANCOVA models were performed. Model terms included CD, HIV, the interaction between CD and HIV (CD*HIV), age, sex, and years of education. *Post-hoc* t-tests were further used to characterize any observed effects in the omnibus ANCOVA analysis. Brain volume data were reported as a percentage of eTIV, computed by dividing the raw volume estimate by the participant’s eTIV and multiplying the resulting value by 100 in order to control for the effect of head size on regional brain volume.

#### 2.10.3 Exploratory Whole-Brain Analysis

As a follow-up to the ROI analysis, an exploratory whole-brain analysis was conducted to identify additional regions showing effects of HIV, a history of CD, or the combination of these factors on volume of subcortical structures and cortical thickness.

For cortical thickness, whole-brain vertex-wise analyses were conducted in Freesurfer. Model terms included the factors CD, HIV, CD*HIV, age, sex, and years of education. Because an inverse relationship between disease state and cortical thickness was hypothesized (Hirsiger et al., 2019), a 1-tailed test was used.

For subcortical regions and macrostructural volume measures, ANCOVAs were conducted using the same model factors. Results were corrected for multiple comparison using the Benjamini-Hochberg procedure (Benjamini and Hochberg, 1995). To control for the effect of head size on brain volume measures, all analyses involving brain volume were computed on data normalized for estimated total intracranial volume (eTIV).

#### 2.10.4 Neurocognitive Testing

To examine between-group differences in neurocognitive function—including working memory, verbal learning, task-switching, and motor functioning —a battery of neurocognitive tests and questionnaires was administered (**Table 2, Figure 2**). A two-phase approach was used to characterize the effect of CD, HIV, and their combination, on neurocognitive test performance. In phase 1, neurocognitive and behavioral testing scores were reported from each test separately **(Table 2**). For assessments reporting age-normalized T-scores as the primary outcome measure (WAIS, Hopkins), results were tested for significance using 2-way ANOVAs with factors CD, HIV, CD*HIV, sex, and years of education. For all other tests in which the primary outcome measure was not age-normalized, age was included as an additional model factor. For Trails A and B data, an “efficiency score” was calculated for each subject by the following formula: Efficiency = (Total Correct Matches – Total Number of Errors) / Completion Time in Seconds.

**Table 2:**
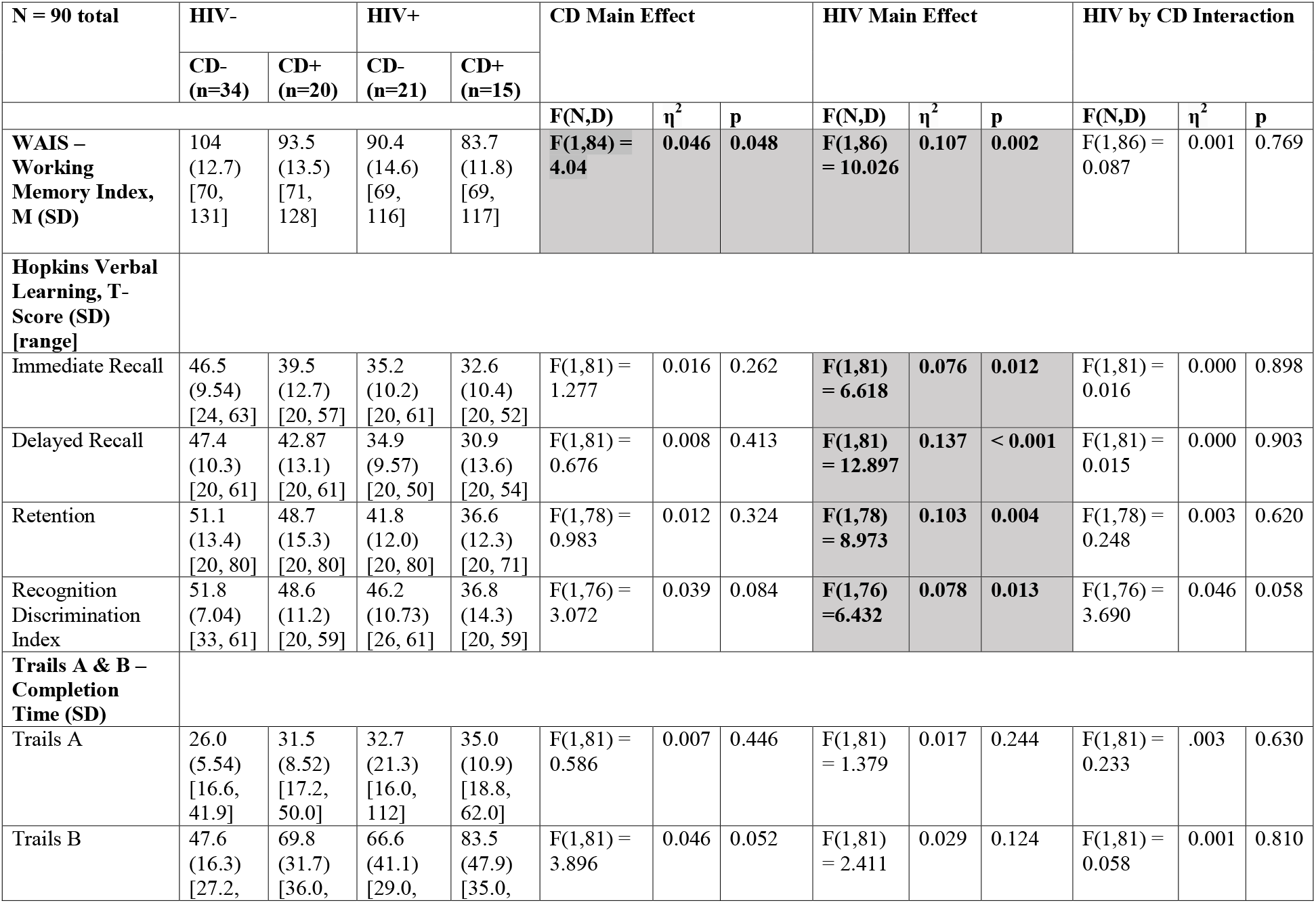
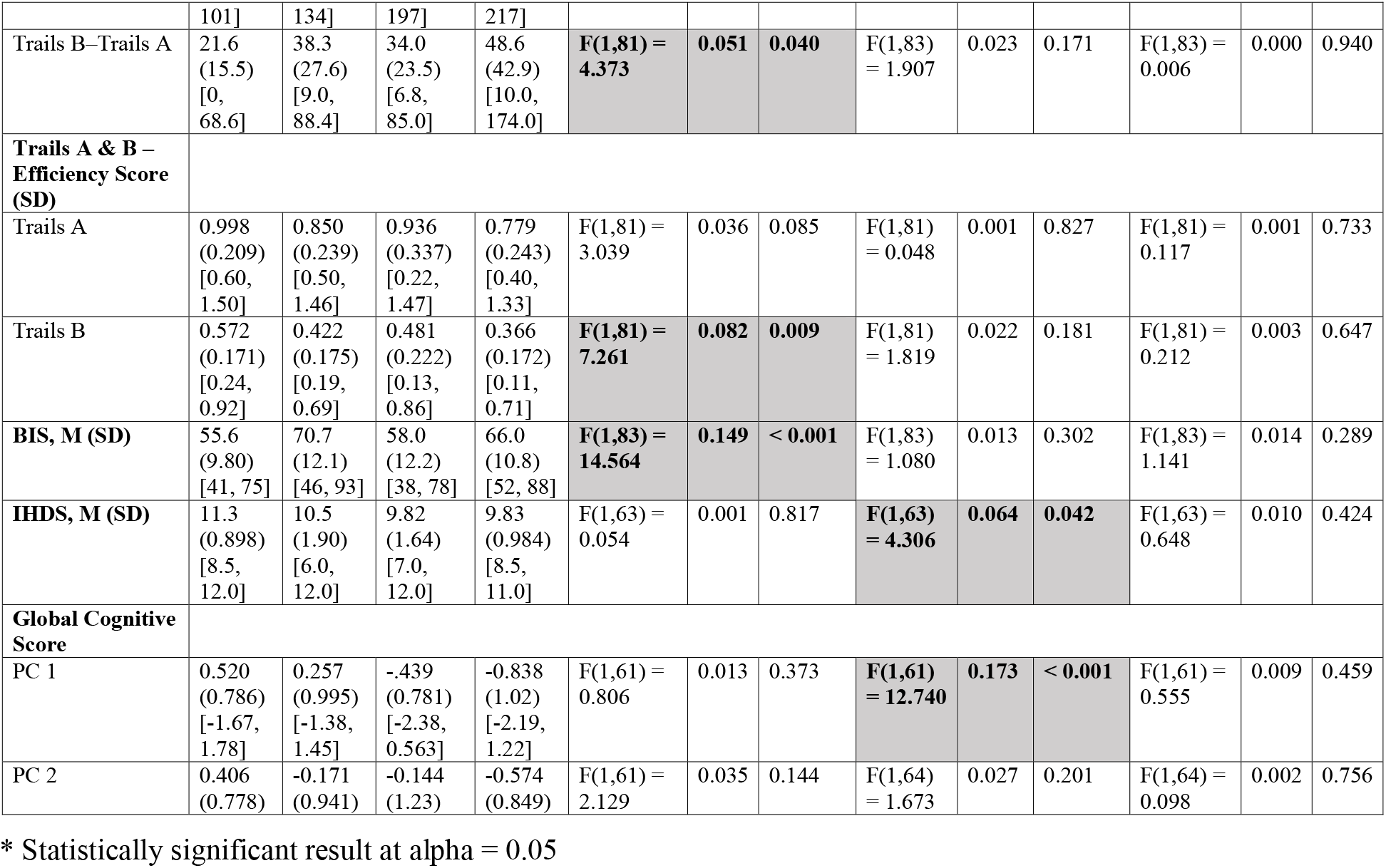
Neurocognitive Assessment Scores. Shown as Mean (SD). Shaded cells indicate statistically significant comparisons. Model factors for assessments reporting age-normalized T-scores as the primary outcome measure (WAIS, Hopkins) were CD, HIV, and CD*HIV, sex, and years of education. Model factors for all other assessments were CD, HIV, CD*HIV, age, sex, and years of education.

**Figure 2:**
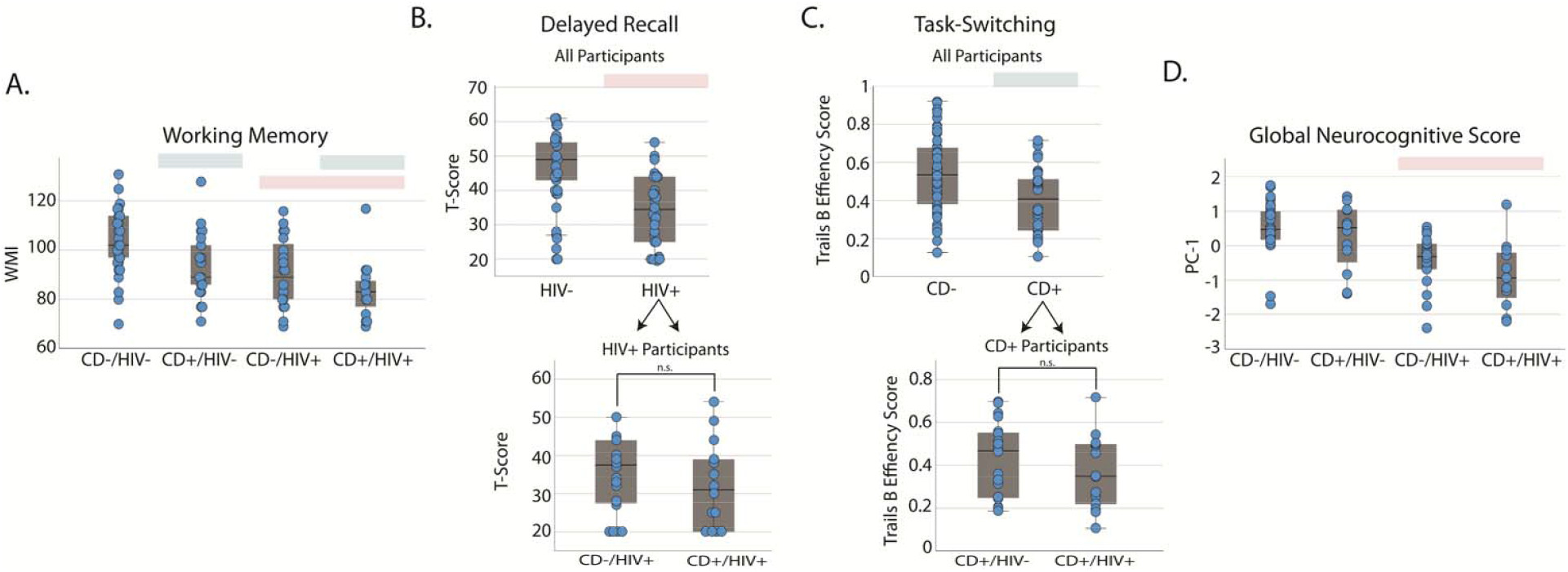
Neurocognitive Assessments. A graphical depiction of a selection of results shown in Table 2.Green shaded bars indicate a significant main effect of CD history, and pink shaded bars indicate a significant main effect of HIV. (A) WAIS – Working Memory Subsection, (B) Hopkins Verbal Learning – Delayed Recall Subsection, (C) Trails B Efficacy Score, (D) Global Neurocognitive Score (PC 1)

In phase 2, a “Global Neurocognitive Score” was computed by combining scores derived from WAIS-WMI, HVLT-R (Immediate, Delayed, RDI, Retention), Trails A and B efficiency scores, IHDS, and BIS using principle component analysis (PCA)-based factor analysis with varimax rotation (Costello and Osborne, 2005). Before PCA, each measure was normalized by subtracting the mean across all subjects and dividing by the standard deviation. Participants with any missing neurocognitive testing data were excluded from this analysis. ANCOVAs with model factors CD, HIV, CD*HIV, age, sex and years of education were performed on data extracted from the first two principle components (PC) in order to investigate possible effects of clinical diagnosis on neurocognitive performance while controlling for demographic variables.

#### 2.9.5 Relationship between Neurocognitive Performance and Cortical Thickness

As an exploratory aim to investigate whether cortical thickness was related to Global Neurocognitive Score at any brain locus, whole-brain vertex-wise comparison between Global Neurocognitive Score (PC-1) and cortical thickness were performed for each vertex of the cortex across all participants. Due to an a priori hypothesis that higher cortical thickness would relate to greater neurocognitive functioning, a 1-tailed test was used to assess statistical significance.

#### 2.9.6 Relationship between Clinical Measures of Disease Severity and Cortical Thickness

To investigate the relationship between cortical thickness and clinical measures of disease severity including duration of abstinence, duration of cocaine usage, and CD4-count, whole-brain vertex-wise correlations were performed. To account for the effect demographic variables, age, sex and years of education were included as model covariates. Four CD+ outliers (2 CD+/HIV-, 2 CD+/HIV+) were excluded from analyses involving duration of cocaine abstinence due to durations of cocaine abstinence greater than 3 years. One CD-/HIV+ outlier was excluded from correlation analyses involving current CD4 count due to a documented CD4 count outside of the physiological range (3008 cells/mm^3^) and greater than 5 STD from the mean of all other HIV+ participants. Because an inverse relationship between cortical thickness and measures of disease severity was hypothesized (Hirsiger et al., 2019; Sanford et al., 2018; Sanford et al., 2017), a 1-tailed test was used to assess statistical significance.

## 3 Results

### 3.1 Participants

Participants across groups were comparable in age, biological sex, and number of persons testing positive for THC on the date of the MRI scan **(Table 1)**. There were no significant between-group differences in HIV-related clinical measures (viral load, current CD4-count, duration of HIV infection), and no differences between CD+ groups in CD-related clinical measures (lifetime cocaine usage, duration of cocaine abstinence). Although CD+ groups were comparable on past cocaine, alcohol, and tobacco use severity, CD+/HIV-participants reported more severe patterns of past opiate usage compared to CD+/HIV+ participants.

Across all groups, significant differences were observed in group racial composition, years of education, number of THC positive urine samples on the date of neurocognitive testing, and current smoking status. Both CD+ groups were comparable on current smoking status, and CD+ participants were significantly more likely to identify as current smokers compared to CD-participants.

### 3.2 Neurocognitive Testing

After controlling for demographic variables, main effects of either CD, HIV or both CD and HIV were observed across all assessments. Significant main effects for both CD and HIV were seen on assessments of working memory (HIV: F(1,84) = 10.026, η^2^ = 0.107, p = 0.002; CD: (F(1,84) = 4.04, η^2^ = 0.0.046, p = 0.048) (**Table 2, Figure 2A)**. For assessments of verbal learning, significant effects of HIV were present across all HVLT subsections (**Table 2, Figure 2B)**. In addition, HIV+ status was associated with significantly reduced performance on the International HIV Dementia Test (IHDS) (F(1,63) = 4.306, η^2^ = 0.064, p = 0.042). On the Trails B assessment of task-switching, CD participants showed significantly lower efficacy (F(1,81) = 7.261, η^2^ = 0.082, p = 0.009) and slower performance (F(1,81) = 4.373, η^2^ = 0.051, 0.040) compared to CD-participants (**Table 2, Figure 2C**). CD participants also endorsed significantly higher impulsivity on the BIS questionnaire (F(1,83) = 14.564, η^2^ =0.149, p < 0.001) (**Table 2**). Although no significant interactions between CD history and HIV were observed, CD+/HIV+ patients demonstrated on average the lowest performance across all neurocognitive assessments. PCA-based factor analysis indicated that PC 1 explained 50.638% of the variance within the data (**Figure 2D, Table 2**). After controlling for participant age, sex, and years of education, a main effect of HIV along this component was identified (F(1,61) = 12.740, η^2^ = 0.173, p < 0.001), indicating that HIV status is a primary driver of reduced neurocognitive performance (**Figure 2D)**. PC 2 was found to explain 13.562% of the variance in the data. The ANCOVA analysis on PC 2 data revealed no significant effect of any factor of interest (CD, HIV, or CD*HIV) (**Table 2**) or demographic covariate.

### 3.3 Brain Morphometry

To examine the relationship between CD history, HIV, and macrostructral brain measures, ANCOVAs were performed across ROI volume (caudate, putamen, insula, ACC, OFC) and cortical thickness (insula, ACC, OFC) (**Table 3**). A significant main effect of CD history was observed in the bilateral caudate (F(1,79) = 5.916, η^2^ = 0.070, p = 0.017), indicating higher volume in CD+ participants relative to CD-participants (**Figure 3A**). *Post-hoc* t-tests indicated no significant difference in caudate volume between CD+/HIV-participants and CD+/HIV+ participants (t(32)= -0.986, D = -0.344, p = 0.332) (**Figure 3B**).

**Table 3:** Brain Morphometry. Shown as Mean (SD). Shaded cells indicate statistically significant comparisons. Model factors were CD, HIV, CD*HIV, age, sex, and years of education.

| N = 86 | **HIV-** |  | **HIV+** |  | **CD Main Effect** |  |  | **HIV Main Effect** |  |  | **HIV by CD Interaction** |  |  |
| --- | --- | --- | --- | --- | --- | --- | --- | --- | --- | --- | --- | --- | --- |
|  | **CD-**<br>**(n=32)** | **CD+**<br>**(n=20)** | **CD-**<br>**(n=20)** | **CD+**<br>**(n=14)** |  |  |  |  |  |  |  |  |  |
|  |  |  |  |  | **F(1,81)** | **$\eta^2$** | **p** | **F(1,81)** | **$\eta^2$** | **p** | **F(1,81)** | **$\eta^2$** | **p** |
| **Volume (% of eTIV)** |  |  |  |  |  |  |  |  |  |  |  |  |  |
| **Caudate** | 0.464<br>(0.054) | 0.482<br>(0.067) | 0.461<br>(0.062) | 0.506<br>(0.076) | **5.916** | **0.070** | **0.017\*** | 1.852 | 0.023 | 0.177 | 0.958 | 0.012 | 0.331 |
| **Putamen** | 0.651<br>(0.090) | 0.683<br>(0.069) | 0.670<br>(0.071) | 0.668<br>(0.062) | 1.355 | 0.017 | 0.248 | 0.873 | 0.011 | 0.353 | 0.441 | 0.006 | 0.509 |
| **Insula** | 0.953<br>(0.067) | 0.948<br>(0.089) | 0.943<br>(0.112) | 0.953<br>(0.094) | 0.046 | 0.001 | 0.831 | 0.052 | 0.001 | 0.821 | 0.388 | 0.005 | 0.535 |
| **ACC** | 0.304<br>(0.044) | 0.313<br>(0.056) | 0.302<br>(0.043) | 0.294<br>(0.036) | 0.049 | 0.001 | 0.775 | 0.554 | 0.007 | 0.459 | 1.661 | 0.021 | 0.201 |
| **Medial OFC** | 0.692<br>(0.066) | 0.712<br>(0.056) | 0.687<br>(0.061) | 0.676<br>(0.066) | 0.989 | 0.012 | 0.323 | 0.507 | 0.006 | 0.479 | 1.534 | 0.019 | 0.219 |
| **Thickness (mm)** |  |  |  |  |  |  |  |  |  |  |  |  |  |
| **Insula** | 2.919<br>(0.144) | 2.842<br>(0.178) | 2.861<br>(0.183) | 2.798<br>(0.169) | 3.089 | 0.038 | 0.083 | 0.468 | 0.006 | 0.496 | 0.822 | 0.010 | 0.367 |
| **ACC** | 2.752<br>(0.199) | 2.741<br>(0.165) | 2.689<br>(0.209) | 2.645<br>(0.149) | 0.003 | 0.000 | 0.955 | 0.668 | 0.008 | 0.416 | 0.016 | 0.000 | 0.900 |
| **Medial OFC** | 2.356<br>(0.115) | 2.356<br>(0.142) | 2.366<br>(0.122) | 2.291<br>(0.116) | 0.442 | 0.005 | 0.518 | 0.181 | 0.002 | 0.672 | 1.571 | 0.019 | 0.214 |

\* Statistically significant result at $\alpha = 0.05$

**Figure 3:**
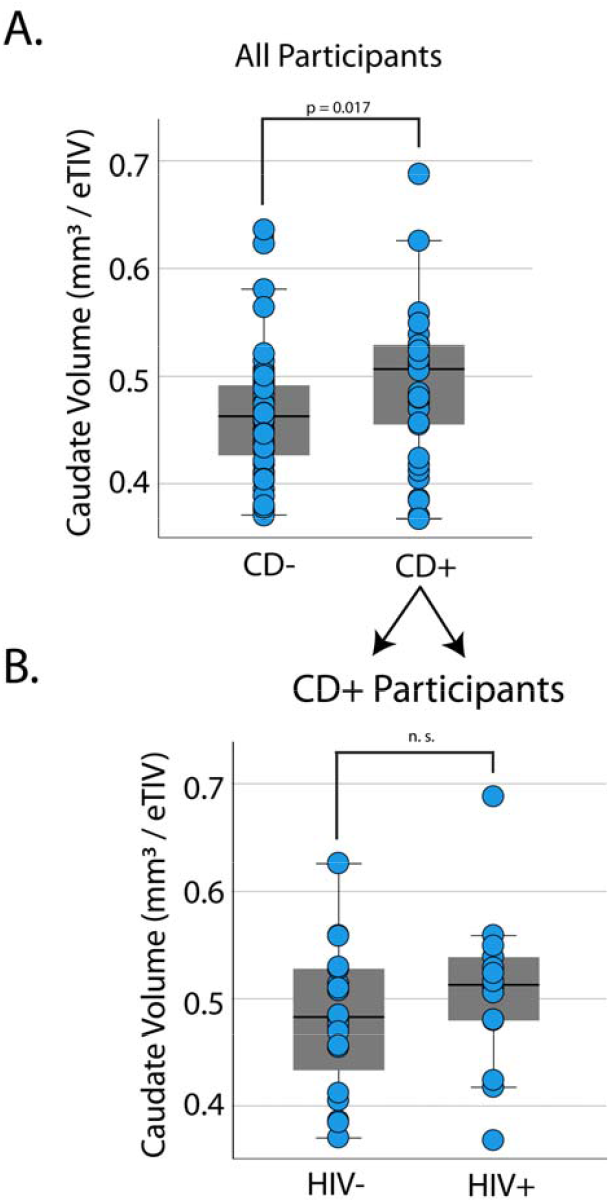
Brain Morphometry. Visualization of the main effect of CD on caudate volume from Table 2. (A) Bilateral caudate volume expressed as percentage of eTIV, plotted separately for CD+ and CD-participants. Dots indicate individual participant values. (B) Bilateral caudate volume for CD+ participants expressed as percentage of eTIV and plotted as a function of HIV.

### 3.4 Relationship Between Cortical Thickness and Neurocognitive Performance

In order to characterize the relationship between cortical thickness and neurocognitive performance, whole-brain correlations were conducted between cortical thickness at each vertex and Global Neurocognitive Score. Following cluster correction, no significant correlations were identified at any cortical locus.

### 3.5 Relationship Between Cortical Thickness and Measures of Disease Severity

As an exploratory aim to investigate possible relationships between measures of clinical disease severity and cortical thickness while controlling for demographic variables, whole-brain vertex-wise GLMs were conducted between cortical thickness and clinical measures, including duration of infection, duration of abstinence, duration of usage, and current CD4 count. Following cluster correction, no significant correlations were identified.

### 3.6 Whole-Brain Vertex-Wise Analysis

To identify additional cortical regions showing differences in macrostructural measures as a function of HIV, CD, or their combination while controlling for demographic variables (age, sex, years of education), whole-brain vertex-wise GLMs were conducted Freesurfer. Following cluster correction, no significant correlations were identified at any cortical locus. Because of the tightly-coupled association between educational attainment, socio-economic status and addiction, analyses were repeated without the inclusion of years of education as a covariate. Results are shown in **Figure S1** for the purpose of future hypothesis generation.

To identify additional possible subcortical regions and macrostructural measures showing possible effects of clinical diagnosis on subcortical volume, 2-way ANCOVAs with factors CD, HIV, CD*HIV, age, sex, and years of education were conducted for each measure. Following Benjamini-Hochberg correction, no regions survived. Results of regions showing effects of clinical diagnosis on volume measures prior to multiple comparison correction are given in **Figure S2** for the purpose of future hypothesis generation, but these results are not interpreted in text.

## Discussion

The purpose of the current study was to investigate the relationship between HIV on macrostructural brain measures and neurocognitive performance in individuals with and without a history of CD. Although CD+/HIV+ participants demonstrated the greatest deviations from healthy control participants in measures of brain integrity and neurocognitive test performance, this study did not find evidence of interactive effects of the combination of CD history and HIV on neurocognitive function and macrostructural brain measures in patients with comorbid HIV and CD history.

Using PCA, HIV was identified as a major driver of cognitive performance declines in patient groups. This finding aligns with a large body of literature indicating persistent neurocognitive impairment in the modern era of antiretroviral medication (Heaton et al., 2010). Surprisingly, no significant effects of HIV on cortical thickness were present after controlling for demographic variables in both a priori-defined regions of interest and in a whole-brain vertex-wise analysis. Further, no significant differences in cortical or subcortical volume were observed as a function of HIV in both the regions of interest and at the whole-brain level. These findings contrast with similar reports indicating macrostructural brain difference between HIV+ and HIV-individuals in both cortical and subcortical region in the post-HAART era (Ances et al., 2012a; Correa et al., 2016; Ragin et al., 2012). While the current findings may provide evidence of increased neuroprotection afforded by early HIV detection as well as modern antiretroviral treatments, these results may also highlight heterogeneities in statistical thresholding between different studies. Recent literature has shown high rates of false-positive cluster detection using conventional thresholding (Greve and Fischl, 2018). As such, the current work employed a cluster-detection threshold of p < 0.005 followed by a cluster-probability threshold of p < 0.05. In accordance with Freesurfer guidelines, an additional Bonferroni correction was applied to account for tests performed across both hemispheres. Many works published prior to 2018 employed more liberal cluster thresholding procedures (Lewis-de Los Angeles et al., 2016; Nwosu et al., 2018), complicating the comparison of findings across different studies. Further, in the current study, all statistical models were designed to control for demographic variables, including educational attainment, a well-established proxy measure for socio-economic status. When analyses were repeated without the inclusion of this covariate, modest cortical atrophy was observed as a function of HIV in the left posterior frontal lobe. Indeed, across all participants included in MRI analyses, HIV+ individuals showed significantly lower educational attainment compared to HIV-individuals (HIV-: 15.44 ± 3.177 years; HIV+: 13.06 ± 2.159; t(84) = 3.830, p < 0.001). It is possible that educational attainment and HIV status overlap in terms of the variance explained by these variables on cortical thickness.

Data from the current study provided preliminary evidence of higher caudate volume CD+ individuals relative to CD-. While it was not possible in the current cross-sectional design to tease apart whether the observed macrostructural differences resulted from cocaine usage or preceded it, high caudate volume may represent a preexisting vulnerability marker for substance use disorder, since higher striatal volume has been observed in non-dependent siblings of patients with CD relative to healthy volunteers (Ersche et al., 2012). As an alternative interpretation, data from animal models suggests that repeated exposure to psychostimulant drugs can lead to increased dendritic branching and spine density in neurons of the basal ganglia (Robinson and Kolb, 1999): a finding which may contribute to the higher caudate volume estimates observed in CD+ participants in the current work. It should also be noted that findings of CD-related striatal volume differences in the literature are generally mixed, with some neuroimaging (Ersche et al., 2012; Jernigan et al., 2005) and animal studies (Wheeler et al., 2013) indicating higher striatal volume as a function of stimulant drug dependence, while others indicate lower striatal volume (Barros-Loscertales et al., 2011). The current data would support the former.

Contrary to the initial hypothesis, the current study found no evidence of an interaction between HIV and history of CD on neurocognitive performance, nor evidence of interactive changes in measures of brain integrity in participants with HIV and history of CD. Although previous functional neuroimaging studies have revealed synergistic CD+/HIV+ alterations in executive control-related brain circuitry (Bauer, 2011; Bell et al., 2020; Meade et al., 2016; Meade et al., 2017; Wakim et al., 2021), only three studies to date have employed structural MRI to examine the possible interactive effects of HIV and substance dependence on brain-based measures using structural neuroimaging. Jernigan and colleagues used structural MRI to examine the interaction between a history of methamphetamine (meth) dependence and HIV in meth-abstinent participants, finding largely independent effects of each factor on cerebral brain volume estimates (Jernigan et al., 2005). Using a similar study design, Macduffie and colleagues examined the interaction between meth dependence and HIV on cortical thickness, area, and volume, finding no evidence that meth modulated brain morphology in any cortical area (MacDuffie et al., 2018). Cordero and colleagues used DTI to examine the effects of comorbid CD and HIV on white matter integrity, with findings again largely indicating independent effects of each factor (Cordero et al., 2017).

There are a number of limitations of the current work. Although participant groups were similar in age, sex, and clinical variables, there were significant between-group differences in race, years of education, and smoking status, which may have impacted the results reported here. For example, recent work has shown that daily cigarette smoking is associated with more rapid age-related cortical thinning, particularly in the anterior frontal and insular cortices (Durazzo et al., 2017): regions which overlap with those of interest in the current work. However, it should be noted that the prevalence of cigarette smoking for patients in treatment for substance use disorders in the United States (76.3%) (Guydish et al., 2011) is markedly higher than in the general population (14% in 2019) (Cornelius et al., 2020), and that the current sample—in which 75% of CD+ participants identified as current smokers, compared to 15% of the CD-sample—therefore represents an accurate reflection of the broader population of individuals in recovery relative to the general population. While there were no significant differences in sex ratios between groups, the CD+/HIV+ healthy control group contained more female members than any other group. In addition, participants in HIV-groups were younger on average than participants in the HIV+ groups (HIV-: 42.7 ± 11.4; HIV+: 46.6 ± 11.1), although this difference does not reach statistical significance. Finally, since data on participant sexual orientation was not collected in the course of the current investigation, it is unknown whether participant groups were matched on sexual orientation: a factor which has been recently linked to gray matter volume differences in subcortical brain regions (Votinov et al., 2021).

In summary, no synergistic changes in brain morphometry or neurocognitive capabilities were observed in patients with comorbid CD and HIV when compared against individuals with either condition alone. Using PCA, HIV was identified as a primary driver of neurocognitive impairment. Higher caudate volume was identified in CD+ participants relative to CD-. All data are available upon reasonable request to principle investigator.

## Data Availability

Data will be made available upon reasonable request to the corresponding author.

## Author Contributions Statement

JJF and KMW were responsible for the initial study concept. KMW was responsible for participant recruitment and data collection. KMW performed the primary data analyses with invaluable assistance of MET and ZC. KMW wrote the first draft of the manuscript. JJF and EGF were site PIs and oversaw all procedures. JJF, EGF MET, and SM provided editorial input and critical revisions of the manuscript. All authors approve of this final version for publication.

## Acknowledgments

We thank the entire team at the Cognitive Neurophysiology Laboratory at the University of Rochester and Albert Einstein College of Medicine for their continued support. We thank Kelly Farrow, N.P, Giovani Schifitto, M.D., and Teresa Oh, B.S., for invaluable patient referrals. We thank Leona Oakes, Ph.D., for clinical phenotyping supervision and consultation.

## Data Sharing

Data will be made available upon reasonable request to the corresponding author.

---

**Figure S1:**
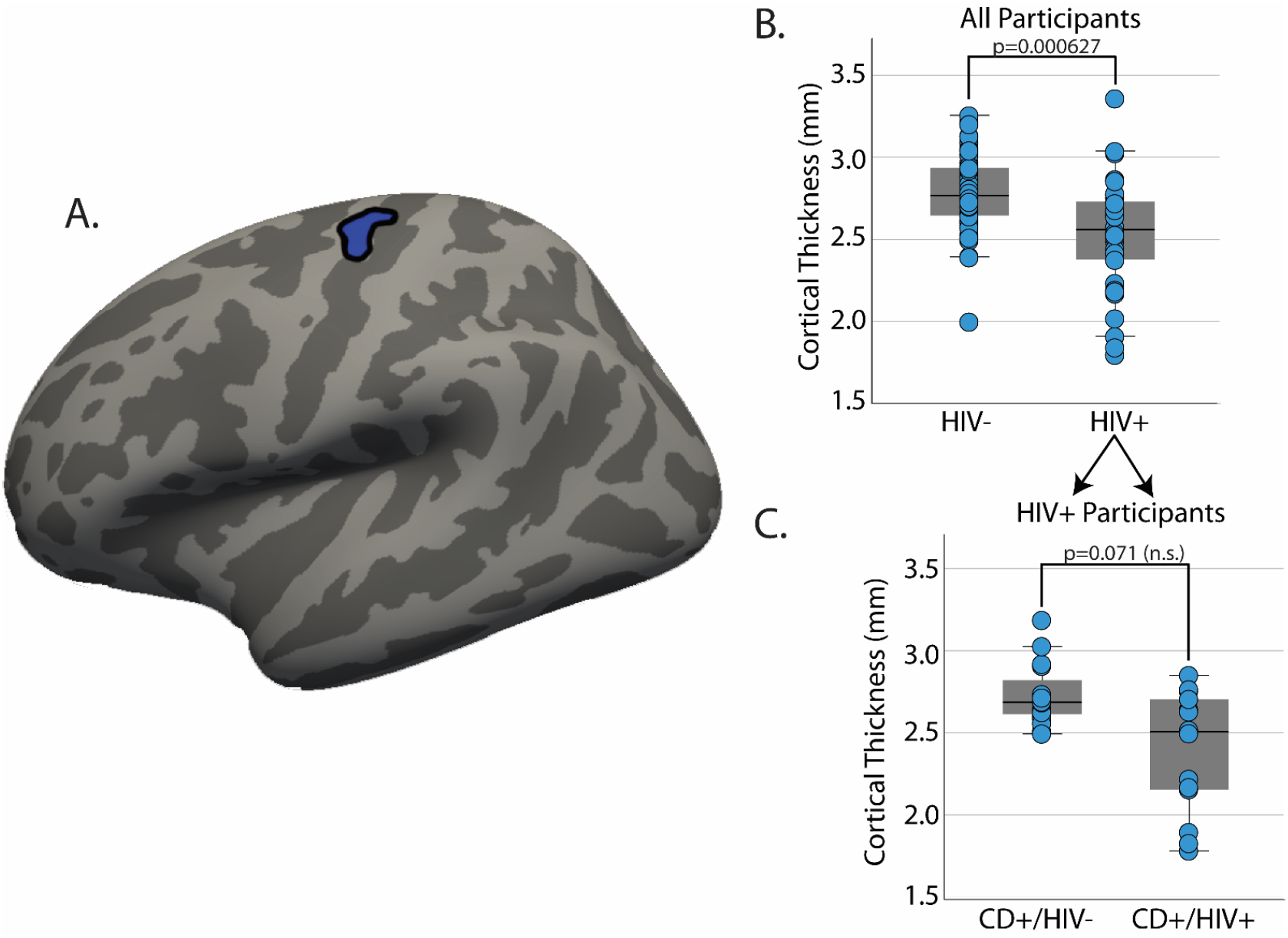
Exploratory Whole-Brain Analysis of Cortical Thickness: Main effects of HIV on cortical thickness were observed in the left prefrontal gyrus. Model factors included CD, HIV, CD*HIV, age, and sex. (A) Left prefrontal gyrus cluster projected in surface space. (B) Visualization of the HIV main effect across all subjects. (C) Visualization of data distribution in HIV participants plotted separately for CD+ participants (right) and CD-participants (left)

**Figure S2:**
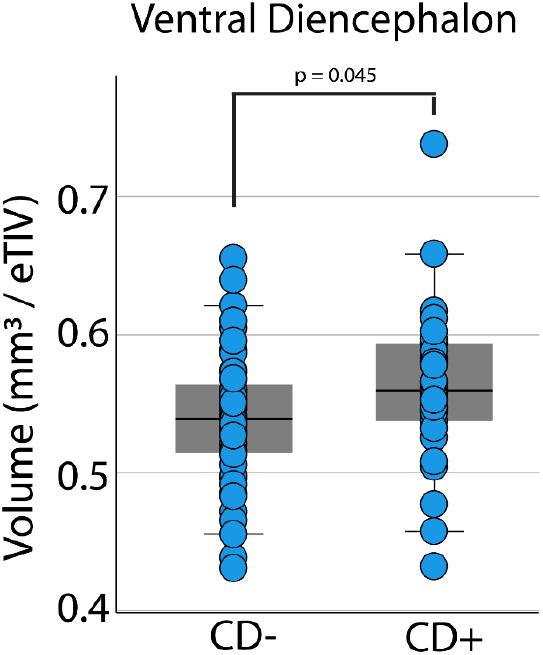
Exploratory Whole-Brain Analysis of Subcortical Volume: A significant effect of main effect CD on brain volume in the ventral diencephalon was observed prior to correction for multiple comparisons in an ANCOVA model controlling for age, sex, and years of education.

**Table S1:** Demographics of MRI Sample. Characteristics by study group for participants comprising the final sample for analysis of MRI data.

| n = 86 Participants | HIV- |  | HIV+ |  | Statistic | Effect Size | *p* value |
| --- | --- | --- | --- | --- | --- | --- | --- |
|  | CD- (n = 32) | CD+ (n = 20) | CD- (n = 20) | CD+ (n = 14) |  |  |  |
| Demographic Characteristics |  |  |  |  |  |  |  |
| Sex Assigned at Birth (N Male : N Female) | 12 : 20 | 11 : 9 | 13<sup>#</sup> : 7 | 7 : 7 | $\chi^2(3) = 4.000$ | $\Phi = 0.216$ | 0.261 |
| Age in years, M (SD) [range] | 42.3 (12.6) [19.3, 65.4] | 42.3 (10.4) [26.0, 61.1] | 44.7 (13.1) [22.9, 64.8] | 48.9 (7.6) [31.8, 58.4] | $F(3,82) = 1.236$ | $\eta^2 = 0.043$ | 0.302 |
| Race, N | | | | | $\chi^2(6) = 21.164$ | $\Phi = 0.496$ | 0.001* |
| African American | 3 | 6 | 9 | 9 |  |  |  |
| Caucasian | 23 | 14 | 10 | 9 |  |  |  |
| Asian | 6 | 0 | 1 | 0 |  |  |  |
| Education in years, M (SD) [range] | 16.7 (2.81) [12, 24] | 13.5 (2.72) [11, 23] | 13.4 (2.11) [9, 18] | 12.6 (2.24) [10, 16] | $F(3,82) = 12.754$ | $\eta^2 = 0.318$ | < 0.001* |
| THC positive urine, N |  |  |  |  |  |  |  |
| Date of Scan | 3 | 2 | 6 | 2 | $\chi^2(3) = 4.140$ | $\Phi = 0.234$ | 0.218 |
| Date of Cognitive Testing | 2 | 1 | 6 | 2 | $\chi^2(3) = 6.524$ | $\Phi = 0.298$ | 0.061 |
| Drug use characteristics |  |  |  |  |  |  |  |
| Current Smoker, N | 2 | 16 | 5 | 10 | $\chi^2(3) = 36.600$ | $\Phi = 0.652$ | < 0.001* |
| **Lifetime cocaine usage in years, M (SD)** | | 14.2 (10.6)<br>[1, 35] | | 19.8 (9.04)<br>[7, 33] | $t(32) = -1.629$ | $D^S = -0.568$ | 0.113 |
| **Duration of abstinence, M (SD)** | | 1.07 (1.03)<br>[0.04, 4.82] | | 1.10 (1.38)<br>[0.07, 4.99] | $t(32) = -0.076$ | $D^S = -0.026$ | 0.094 |
| **HIV Characteristics** |  |  |  |  |  |  |  |
| **HIV viral load <20 copies, N (%)** | | | 20 (100%) | 12 (85.7%) | $\chi^2(1) = 3.036$ | $\Phi = -0.299$ | 0.162 |
| **Current CD4 cell count, M (SD) [range]** | | | 760 (589)<br>[301, 3008] | 730 (236) [190, 1004] | $t(32) = 0.174$ | $D^S = 0.061$ | 0.863 |
| **Years since diagnosis, M (SD) [range]** | | | 16.9 (11.5)<br>[0.80, 34.0] | 16.0 (7.61)<br>[7.57, 33.7] | $t(31.978) = 0.164^{\wedge}$ | $D^S = 0.053$ | 0.871 |

